# The impact of the COVID19 pandemic and initial period of lockdown on the mental health and wellbeing of UK adults

**DOI:** 10.1101/2020.04.24.20078550

**Authors:** Ross G. White, Carine Van der Boor

## Abstract

Mental health and wellbeing impacts of COVID19 were assessed in a convenience sample of 600 UK adults using a cross-sectional design. Recruited over a two-week period during the initial phase of the government lockdown, participants completed an online survey that included COVID19-related questions, the Hospital Anxiety and Depression Scale, the WHO-5 and the OXCAP-MH. Self-isolating prior to lockdown, increased feelings of isolation since the lockdown, and having COVID19-related livelihood concerns, were associated with poorer mental health, wellbeing and quality of life. Perceiving increased kindness, community connectedness, and being an essential worker were associated with better mental health and wellbeing outcomes.

## Introduction

On 24^th^ March 2020 the UK introduced a range of ‘lockdown’ restrictions intended to slow the progression of the COVID19 outbreak. Emerging evidence indicates that the COVID19 outbreak is associated with adverse mental health and wellbeing outcomes for healthcare workers in China^1^ and the general public in Italy^2^. The current study investigated whether mental health and wellbeing outcomes in UK adults are associated with: 1) Experiencing symptoms of COVID19; 2) Being in a group vulnerable to COVID19; 3) Being categorised as an ‘essential worker’; 4) Experiencing COVID19-related isolation; 5) Local community interactions. Furthermore, the current study explored if level of social support was associated with mental health and wellbeing outcomes.

## Methods

A cross-sectional design was used. A convenience sample recruited via social media completed an online survey. Data was collected over 2-weeks in the initial lockdown period (31^st^ March to 13^th^ April 2020). To be eligible, people had to be adults (+18 years), speak English, and be living in the UK at the time of the COVID19 outbreak. The authors assert that all procedures contributing to this work comply with the ethical standards of the relevant national and institutional committees on human experimentation and with the Helsinki Declaration of 1975, as revised in 2008. All procedures involving human participants were approved by the Central University Research Ethics Committees, University of Liverpool (ref:7633). Written informed consent was obtained from all participants.

The survey included: (i) demographic questions, (ii) COVID19-related questions, (iii) Hospital Anxiety and Depression Scale (HADS^3^), (iv) WHO-5 measure of well-being^4^, (v) Oxford CAPabilities questionnaire-Mental Health (OXCAP-MH^5^) measure of quality of life (QoL), (vi) Multidimensional Scale of Perceived Social Support^6^.

Six-hundred participants (74% female, mean age = 36.75 years, SD = 13.44, range = 18-76 years) completed at a minimum the demographic and COVID19-related questions. Participants were mainly White (93.6%) and employed (65%). Around a quarter of participants (26.3%) self-reported currently receiving treatment for mental disorders – including mood disorders (18%) and neurotic, stress-related and somatoform disorders (14.3%). No participants had been diagnosed with COVID19.

## Results

The mean scores on HADS Anxiety subscale (M=10.23, SD=4.98) and HADS Depression subscale (M=7.57, SD = 4.39) exceeded the “normal” range (i.e. scores of 0-7). The mean scores on WHO-5 and OXCAP-MH were 10.43 (SD = 5.40) and 69.45 (SD = 11.91) respectively.

**Table 1.** Between group analyses for HADS, OXCAP-MH and WHO-5

| Question | Response | Mean | SD | t-value | df | p |
| --- | --- | --- | --- | --- | --- | --- |
| Being in a 'vulnerable group' |  |  |  |  |  |  |
| HADS-Depression | Yes | 7.80 | 4.65 | .48 | 549 | 0.629 |
|  | No | 7.53 | 4.36 |  |  |  |
| HADS-Anxiety | Yes | 9.81 | 5.15 | -.75 | 546 | 0.454 |
|  | No | 10.29 | 4.97 |  |  |  |
| OXCAP-MH | Yes | 68.73 | 13.20 | -.63 | 465 | 0.527 |
|  | No | 69.71 | 11.59 |  |  |  |
| WHO-5 | Yes | 10.44 | 5.83 | .04 | 532 | 0.970 |
|  | No | 10.41 | 5.34 |  |  |  |
| Experienced symptoms of COVID19 |  |  |  |  |  |  |
| HADS-Depression | Yes | 7.68 | 3.40 | .22 | 546 | 0.825 |
|  | No | 7.55 | 4.46 |  |  |  |
| HADS-Anxiety | Yes | 10.56 | 4.74 | .56 | 544 | 0.573 |
|  | No | 10.19 | 5.01 |  |  |  |
| OXCAP-MH | Yes | 66.78 | 14.64 | -1.60 | 65.72 | 0.113 |
|  | No | 70.02 | 11.32 |  |  |  |
| WHO-5 | Yes | 9.82 | 5.46 | -.99 | 530 | 0.322 |
|  | No | 10.52 | 5.40 |  |  |  |
| Self-isolated prior to lockdown due to symptoms of COVID19 |  |  |  |  |  |  |
| HADS-Depression | Yes | 9.00 | 4.36 | 2.83 | 550 | 0.005** |
|  | No | 7.37 | 4.36 |  |  |  |
| HADS-Anxiety | Yes | 11.83 | 4.74 | 2.77 | 548 | 0.006** |
|  | No | 10.02 | 4.98 |  |  |  |
| OXCAP-MH | Yes | 64.42 | 13.25 | -3.56 | 466 | <0.001*** |
|  | No | 70.28 | 11.43 |  |  |  |
| WHO-5 | Yes | 8.98 | 5.29 | -2.29 | 534 | 0.022* |
|  | No | 10.63 | 5.39 |  |  |  |
| Agree felt more isolated than usual during lockdown |  |  |  |  |  |  |
| HADS-Depression | Yes | 8.20 | 4.31 | -7.77 | 250.86 | <0.001*** |
|  | No | 5.16 | 3.68 |  |  |  |
| HADS-Anxiety | Yes | 10.91 | 4.69 | -5.95 | 513 | <0.001*** |
|  | No | 7.99 | 5.14 |  |  |  |
| OXCAP-MH | Yes | 68.53 | 11.46 | 4.16 | 441 | <0.001*** |
|  | No | 73.67 | 11.00 |  |  |  |
| WHO-5 | Yes | 9.64 | 5.07 | 6.18 | 191.84 | <0.001*** |
|  | No | 13.17 | 5.67 |  |  |  |
| Participants identified as an essential worker |  |  |  |  |  |  |
| HADS-Depression | Yes | 7.01 | 4.04 | -2.18 | 400.76 | 0.030* |
|  | No | 7.84 | 4.54 |  |  |  |
| HADS-Anxiety | Yes | 9.83 | 4.79 | -1.30 | 546 | 0.194 |
|  | No | 10.42 | 5.08 |  |  |  |
| OXCAP-MH | Yes | 70.30 | 11.40 | 0.97 | 465 | 0.332 |
|  | No | 69.18 | 12.02 |  |  |  |
| WHO-5 | Yes | 10.82 | 5.12 | 1.18 | 532 | 0.238 |
|  | No | 10.24 | 5.54 |  |  |  |
| Agree that the COVID19 outbreak was threatening their livelihood |  |  |  |  |  |  |
| HADS-Depression | Yes | 8.08 | 4.49 | -2.55 | 544 | 0.011* |
|  | No | 7.13 | 4.23 |  |  |  |
| HADS-Anxiety | Yes | 10.67 | 4.93 | -1.90 | 542 | 0.058 |
|  | No | 9.86 | 4.96 |  |  |  |
| OXCAP-MH | Yes | 68.05 | 11.92 | 2.73 | 461 | 0.007** |
|  | No | 71.02 | 11.47 |  |  |  |
| WHO-5 | Yes | 10.20 | 5.39 | 0.86 | 528 | 0.391 |
|  | No | 10.61 | 5.39 |  |  |  |
| Agree that people's kindness towards others in their local area had increased |  |  |  |  |  |  |
| HADS-Depression | Yes | 7.29 | 4.22 | 2.25 | 551 | 0.025* |
|  | No | 8.20 | 4.70 |  |  |  |
| HADS-Anxiety | Yes | 10.13 | 4.84 | 0.75 | 548 | 0.455 |
|  | No | 10.47 | 5.29 |  |  |  |
| OXCAP-MH | Yes | 71.09 | 11.12 | -4.56 | 467 | <0.001*** |
|  | No | 65.74 | 12.75 |  |  |  |
| WHO-5 | Yes | 10.85 | 5.24 | -2.85 | 535 | 0.005** |
|  | No | 9.42 | 5.62 |  |  |  |
| Agree that since the COVID19 outbreak commenced felt more connected to the members of their local community |  |  |  |  |  |  |
| HADS-Depression | Yes | 7.07 | 4.08 | 2.11 | 552 | 0.035* |
|  | No | 7.87 | 4.56 |  |  |  |
| HADS-Anxiety | Yes | 10.00 | 4.87 | 0.85 | 549 | 0.395 |
|  | No | 10.37 | 5.05 |  |  |  |
| OXCAP-MH | Yes | 72.00 | 10.02 | -3.87 | 467 | <0.001*** |
|  | No | 67.76 | 12.74 |  |  |  |
| WHO-5 | Yes | 11.24 | 5.06 | -2.83 | 536 | 0.005** |
|  | No | 9.90 | 5.55 |  |  |  |

\*p < 0.05; \*\*p < 0.01; \*\*\*p < 0.001

Being in a vulnerable group (12.5%) or experiencing symptoms of COVID19 (11.7%), was not associated with significant differences in mental health and wellbeing outcomes. Participants who self-isolated prior to lockdown due to symptoms of COVID19 (11.8%), relative to those who did not, had higher levels of anxiety (t (584) = 2.77, p = 0.006) and depression (t (550) = 2.83, p = 0.005) and lower levels of wellbeing (t (534) = -2.29, p = 0.022) and QoL (t (466) = -3.56, p < 0.001). Participants who felt more isolated than usual during lockdown (69%) had higher levels of anxiety (t (513)=-5.95, p < 0.001) and depression (t (250.86) = -7.77, p < 0.001), and lower levels of wellbeing (t (191.84) = 6.18, p < 0.001) and QoL (t (441) = 4.16, p < 0.001).

Participants who were essential workers (32%) had significantly lower depression (t (400.76) = -2.18, p = 0.030). Participants who agreed that the COVID19 outbreak was threatening their livelihood (46.0%) had higher depression (t (544) = -2.55, p = 0.011) and lower QoL (t (461) = 2.73, p = 0.007).

Participants who agreed that since the COVID19 outbreak people’s kindness towards others in their local area had increased (68.8%) had lower depression (t (551) = 2.25, p = 0.025), higher QoL (t (467) = -4.56, p < 0.001) and higher wellbeing (t (535)= -2.85, p = 0.005). Similarly, participants who agreed that since the COVID19 outbreak felt more connected to the members of their local community (40.0%) had lower depression (t (552)= 2.11, p = 0.035), and higher QoL (t (467) = -3.87, p < 0.001) and higher wellbeing (t (536)= -2.83, p = 0.005).

The level of perceived social support had significant negative correlations with depression (r = -0.33, p < 0.001) and anxiety (r = -0.17, p < 0.001), and significant positive correlations with QoL (r = 0.52, p < 0.001) and wellbeing (r = 0.29, p < 0.001).

## Discussion

This study sought to investigate the mental health and wellbeing impact of the COVID19 outbreak on a convenience sample of UK adults. The anxiety and depression scores for the sample were markedly higher than normative data derived for the UK adult population’s levels of anxiety (females = 6.78, SD = 4.23; males = 5.51, SD = 4.04) and depression (females = 4.12, SD = 3.78; males = 3.83, SD = 3.74)^7^.

Higher depression scores were associated with participants having to self-isolate prior to lockdown due to symptoms of COVID19, feeling more isolated than usual during lockdown, or agreeing that the COVID19 was threatening their livelihood. On the other hand, agreeing that people’s kindness towards others had increased, agreeing that he/she felt more connected to people in the local community, or working in an essential job was associated with significantly lower depression scores. Notably, the mean depression score for the essential workers (M=7.01, SD = 4.04) remained at the upper limit of the normal range. These findings are open to interpretation, but it may be that the importance of their work, and/or public acknowledgment of their efforts buffer against higher levels of depression.

The significant findings relating to isolation (self-isolating prior to lockdown or feeling more isolated during lockdown) and levels of perceived social support, highlight the importance of exploring innovative ways to maintain connection and social support during periods of lockdown and beyond.

Comparatively high levels of both wellbeing and QoL were associated with participants agreeing that: levels of kindness in the local area had increased, and they felt more connected to others in the local community during the COVID19. QoL, but not wellbeing, was comparatively lower in participants who indicated that their livelihood was threatened by COVID19. We believe that the Capability Approach (Nussbaum 2011), which focuses specifically on the extent to which people have the freedom to engage in valued forms of being and doing (assessed by the OXCAP-MH), provides an important framework for understanding the impact of COVID19 and associated lockdown. There were a number of important limitations associated with the current study. The convenience sample relied on people who access online social media forums. Males and BAME community members were comparatively under-represented in the sample. The cross-sectional nature of the analyses limits the conclusions that can be drawn. This study is part of a longer programme of research aimed at tracking the impact of the COVID19 outbreak and associated government restrictions on mental health and wellbeing.

The study highlights that whereas personal experience of COVID19 symptoms and being part of a group vulnerable to the effects of COVID19 were not associated with poorer mental health and wellbeing, factors associated with isolation and COVID19-related livelihood concerns were. On the other hand, perceiving increased kindness and connectedness in local areas was associated with better mental health and wellbeing outcomes. Further research aimed at mitigating the mental health and wellbeing impacts of public health emergencies is required.

## Data Availability

The data is available from the authors on request.

## Author contribution

Ross G. White formulated the research questions, designed the study, conducted the study, analysed the data and wrote the article.

Carine van der Boor formulated the research questions, designed the study, conducted the study, analysed the data and wrote the article.

## References

1. Lai J, Ma, S, Wang Y, Cai Z, Hu J, Wei N, et al. Factors Associated with Mental Health Outcomes Among Health Care Workers Exposed to Coronavirus Disease 2019. JAMA Network Open 2020; 3(3): e203976.

2. Rossi R, Socci V, Talevi D, Mensi S, Niolu C, Pacitti F, Di Marco A, et al. COVID19 pandemic and lockdown measures impact on mental health among the general population in Italy. medRxiv 2020 https://www.medrxiv.org/content/10.1101/2020.04.09.20057802v1.article-info

3. Zigmond AS & Snaith RP. The Hospital Anxiety and Depression Scale. Acta Psychiatrica Scandinavica 1983; 67(6): 361–370.

4. World Health Organization: Regional Office for Europe. Well-Being measures in primary health care: The DepCare Project. Consensus meeting, Stockholm, 1998.

5. Simon J, Anand P, Gray A, Rugkasa J, Yeeles K, Burns T. Operationalising the capability approach for outcome measurement in mental health research. Soc Sci Med. 2020; 98: 187–96.

6. Zimet GD, Dahlem NW, Zimet SG, Farley GK (1988). The Multidimensional Scale of Perceived Social Support. Journal of Personality Assessment 1988; 52: 30–41.

7. Breeman S, Cotton S, Fielding S, & Jones GT. Normative data for the hospital anxiety and depression scale. Quality of Life Research 2015; 24(2): 391–398.

8. Nussbaum M. Creating capabilities: the human development approach. US: Harvard University Press 2011.

